# Quantification of Subtype Purity in Luminal A Breast Cancer Predicts Clinical Characteristics and Survival

**DOI:** 10.1101/2023.02.27.23286511

**Authors:** Neeraj Kumar, Peter H. Gann, Stephanie M. McGregor, Amit Sethi

**Affiliations:** Alberta Machine Intelligence Institute, Edmonton, Alberta, CANADA; Department of Pathology, College of Medicine, University of Illinois at Chicago University of Illinois Cancer Center, Chicago, IL, USA; University of Wisconsin-Madison, Department of Pathology and Laboratory Medicine University of Wisconsin Carbone Cancer Center, Madison, WI, USA; Department of Electrical Engineering, Indian Institute of Technology Bombay, Mumbai, INDIA

**Author notes:** <u>Corresponding author:</u>* Amit Sethi, PhD.

**Keywords:** breast cancer, intratumor heterogeneity, subtype admixture, matrix factorization

## Abstract

**Purpose:** PAM50 profiling assigns each breast cancer to a single intrinsic subtype based on a bulk tissue sample. However, individual cancers may show evidence of admixture with an alternate subtype that could affect prognosis and treatment response. We developed a method to model subtype admixture using semi-supervised non-negative matrix factorization (ssNMF) of whole transcriptome data and associated it with tumor, molecular, and survival characteristics for Luminal A (LumA) samples.

**Methods:** We combined TCGA and METABRIC cohorts and obtained transcriptome, molecular, and clinical data, which yielded 11,379 gene transcripts in common, and 1,179 cases assigned to LumA. We used ssNMF to compute the subtype admixture proportions of the four major subtypes – pLumA, pLumB, pHER2 and pBasal – for each case and measured associations with tumor characteristics, molecular features, and survival.

**Results:** Luminal A cases with low pLumA transcriptomic proportion were likelier to have non-luminal pathology, higher clinical and genomic risk factors, and lower overall survival (log rank *P* < 10^−5^), independent of age, stage, and tumor size. We found positive associations between pHER2 and HER2-positivity by IHC or FISH; between pLumB and PR negativity; and between pBasal and younger age, node positivity, *TP53* mutation, and EGFR expression. Predominant basal admixture, in contrast to predominant LumB or HER2 admixture, was not associated with shorter survival.

**Conclusions:** Bulk sampling for genomic analyses provides an opportunity to expose intratumor heterogeneity, as reflected by subtype admixture. Our results elucidate the striking extent of diversity among LumA cancers and suggest that determining the extent and type of admixture holds promise for refining individualized therapy. LumA cancers with a high degree of basal admixture appear to have distinct biological characterstics that warrant further study.

## INTRODUCTION

Intrinsic subtyping by PAM50 profiling identifies distinct categories of breast cancer that differ in their tumor characteristics and behavior, while relying on gene expression in a bulk tissue sample. However, individual cancers vary in their adherence to a single prototype, and some might show evidence of admixture with an alternate subtype due to intratumor heterogeneity. Such admixture could affect prognosis and treatment response.

In a previous study, we used expression data for genes included in the PAM50 panel to develop a new metric, Distance Ratio Criteria (DRC), based on the ratio of Mahalanobis distance of a Luminal A (LumA) case from its assigned centroid to the nearest alternate subtype centroid [1; 2]. We showed that this metric could subdivide LumA cases according to purity of the LumA signature and thus identify distinct clinicopathological, molecular, and survival features based on the degree of subtype admixture. We focused on LumA cancers because admixture of this most favorable subtype with any other subtype could be presumed to worsen prognosis.

Here we significantly extend previous work by using semi-supervised non-negative factorization (ssNMF) on whole transcriptome data from a merged METABRIC/TCGA cohort of LumA cases to compute the degree of resemblance of an individual case to each of the four major breast cancer subtypes. Apart from gaining resolution due to the analysis of many more genes, the expanded cohort provides greater statistical power and allows us to explore the attributes of LumA cancers according to their most likely alternate subtype.

## METHODS

### Study populations

We merged two publicly available breast cancer cohorts – Molecular Taxonomy of Breast Cancer International Consortium (METABRIC) cohort and The Cancer Genome Atlas (TCGA) BRCA provisional cohort – downloaded from cBioportal [3]. Data were available from 3,061 total cases, with gene expression measurements for 15,747 overlapping genes, including 1,179 cases assigned to LumA (n = 674 METABRIC and n = 505 TCGA), covering 11,379 genes. We applied data normalization procedures to merge the cohorts; key features of the cohorts and pre-processing steps are presented in **Table S1**.

Previously reported algorithms for intrinsic subtype calls were used to assign each case to one of five PAM50 subtypes (Luminal A, Luminal B, HER2, Basal and Normal) [4]. Re-computed PAM50 classifications were identical to those recorded in the source datasets. We excluded Normal subtype cases from both cohorts, and Claudin-low subtype from METABRIC.

### Semi-supervised non-negative matrix factorization

Non-negative matrix factorization (NMF) is a strong candidate among mathematical techniques to model transcriptomic data as an admixture of underlying metagenes [5]. NMF results can be interpreted as proportions (which cannot be negative) of the underlying components, unlike other popular techniques, such as singular value decomposition. We extended basic NMF by solving an additional optimization problem of linear classification of a patient’s data into one of four PAM50 subtypes simultaneously with the original optimization to minimize the overall data reconstruction error. This dual optimization is called semi-supervised NMF (ssNMF) [6], with the following objective function:

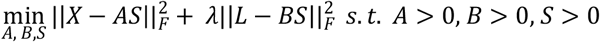

where, *X* ∈ ℝ^*m*×*n*^ represents the matrix containing expression of *m* genes for *n* patients, while the metagenes and their mixing coefficients for factorization rank *k* are given by *A* ∈ ℝ^*m*× *k*^ and *S* ∈ ℝ^*k* × *n*^, respectively, for the data reconstruction error (first) term. Additionally, the one-hot encoded labels for the four PAM50 classes are denoted by matrix *L* ∈ ℝ^4^ × ^*n*^. The basis matrix for the PAM50 label reconstruction term is given by *B* ∈ ℝ^4^× ^*k*^. F represents the Frobenius norm.

In the above stated optimization problem, two hyperparameters need to be set – tradeoff between the two optimization objectives λ > 0, and the number of metagenes *k*. Optimal hyperparameter values for the combined cohort (and for TCGA and METABRIC separately) were obtained based on the accuracy of PAM50 classification using five-fold cross validation.

We interpreted the label reconstruction estimate *BS* ∈ ℝ^4^ × ^*n*^ for the four subtypes as the proportion estimates for subtype admixture. We normalized each column such that all four components (designated pLumA, pLumB, pHER2 and pBasal) sum to one. Since our primary focus was to quantify subtype admixture in PAM50 assigned LumA cases, analyses were performed only on LumA cases. Thus, the proportion of LumA subtype was our primary purity metric for PAM50-assigned Luminal A cases. In the combined cohort, the range of proportions for each subtype was divided into 100 equally spaced intervals to plot histograms. Within the top quartile of each subtype, an exclusive (eQ4) subset was identified, comprising cases not in the top quartile for any other subtype.

### Clinical feature, molecular characteristics and survival analysis

To test the hypothesis that admixed LumA cases had more adverse characteristics than pure ones, we compared clinical and molecular features across quartiles by proportion of LumA transcriptome (pLumA) using two-tailed t-tests or exact chi-square tests. Clinical variables included mean age at diagnosis, percentage with nodal involvement, tumor size > 20mm, stage > I, and ER, PR or HER2 positivity (by immunohistochemistry and/or FISH). Molecular variables evaluated included the PAM50 11-gene Proliferation Score [7], PAM50-based risk of recurrence score [8], Oncotype DX score [9], percent of cases high-risk by MammaPrint [10], and the prevalence of selected somatic mutations. PAM50, Oncotype DX, and MammaPrint scores were all computed from normalized gene expression data using published formulas.

We analyzed the same hypotheses in the individual cohorts, comparing tertiles instead of quartiles for added statistical power. For METABRIC, additional analyses were possible on tumor grade, HER2 copy number gain determined by HER2 SNP6 DNA microarray, proliferation status determined by AURKA expression, and IntClust assignments [11]. For TCGA, additional analyses were done on the number of clonal populations estimated by PyClone, accounting for variations in copy number and benign cell contamination [12].

Overall survival for the combined cohort was analyzed for PAM50-assigned LumA cases stratified by quartiles of pLumA using Kaplan-Meier curves as well as hazard ratios from unadjusted and adjusted Cox proportional hazards models. Factors for adjustment included age, tumor stage and size for combined cohort and TCGA. Similar analyses for METABRIC also adjusted for tumor grade. Associations of tumor characteristics and survival with DRC and Shannon entropy were computed to provide comparison to the results from ssNMF. Reported results are consistent with the REMARK guidelines for prognostic tumor marker studies [13].

### Alternate subtype analysis

Additional analyses compared cases that were exclusively in the highest quartile for pLumA versus those exclusively in the highest quartile for one of the three alternate subtypes. The set eQ4-LumA, representing relatively pure LumA cancers, included 275 cases; eQ4-LumB, eQ4-HER2, and eQ4-Basal comprised 182, 157 and 219 cases, respectively.

## RESULTS

The four proportional distributions, one for each PAM50 class, for the combined TCGA/METABRIC cohort of 1,179 Luminal A cases, are shown in **Figure 1**. Most of the cases had high Luminal A composition with non-zero proportions for other subtypes. The highest quartile for each subtype proportion is highlighted, as is the portion of that quartile comprising cases not in Q4 for any other subtype and thus exclusive Q4.

**Figure 1.**
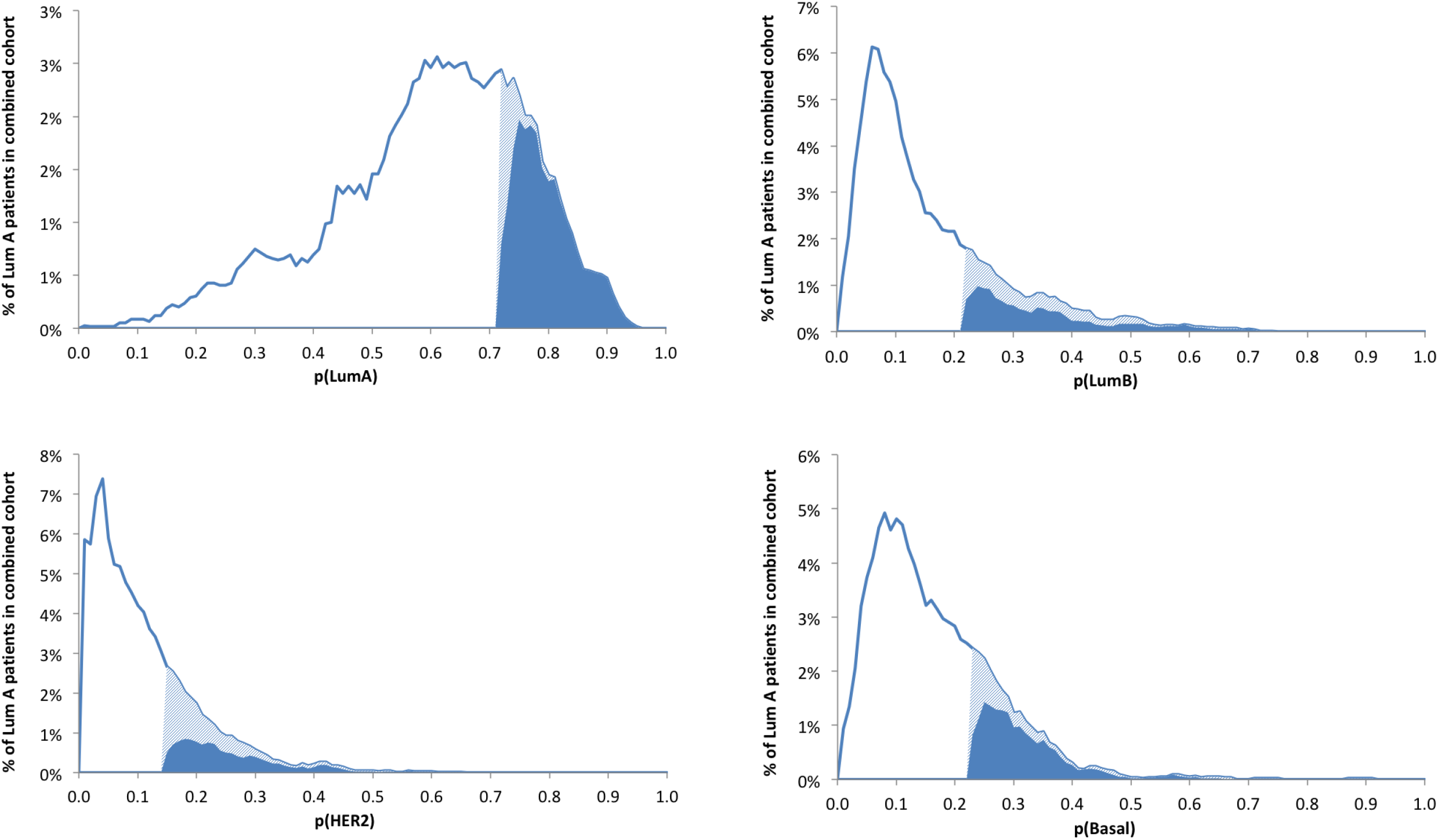
Frequency distributions for proportion of each intrinsic subtype, for all Luminal A breast cancers in combined cohort (TCGA and METABRIC), based on ssNMF analysis of the whole transcriptome. Light plus dark blue shaded area represents the highest quartile; dark blue areas represent the subset of cases that are exclusively in the highest quartile for each alternate subtype.

### Clinical and molecular features in the combined cohort for pLumA

**Table 1** compares clinical-pathological and molecular features of the assigned Luminal A cases according to quartile for pLumA. Compared to relatively “pure” Q4 cases, Q1 cases were on average two years older (*P* trend = 0.027), less likely to be PR-positive, and more likely to be HER2-positive by IHC or FISH. In contrast, purity was not associated with an increase in ER-positivity. However, more Q4 cases adhered to a surrogate definition of Luminal A subtype (ER+ or PR+ and HER2− by IHC) compared to Q1 cases. Triple negative cases (n = 108) were too sparse to permit meaningful conclusions. Q1 status was significantly associated with higher prevalence of lymph node involvement, higher stage, and larger tumor size. These most admixed cases had substantially higher PAM50 proliferation and recurrence scores, and substantially worse scores for both Oncotype DX and MammaPrint gene panels. As pLumA increased, the prevalence of *TP53* mutation decreased almost three-fold, and mutation of *PIK3CA* and *CBFB* - two gene alterations associated with LumA subtype - increased significantly.

**Table 1.** Characteristics of Luminal A breast cancers in the combined cohort (TCGA, METABRIC), stratified by quartile of pLumA subtype purity based on transcriptome

|  | **Q1**<br>n = 295 | **Q2**<br>n = 294 | **Q3**<br>n = 295 | **Q4**<br>n = 295 | **P, Q1 vs Q4**<br>**(p trend)** |
| --- | --- | --- | --- | --- | --- |
| Age (mean) | 62.53 | 61.89 | 60.59 | 60.46 | 0.058<br>(0.027) |
| ER+<sup>a</sup> (%) | 97.23 | 97.89 | 97.93 | 99.30 | 0.106<br>(0.087) |
| PR+ (%) | 76.22 | 82.07 | 86.21 | 84.83 | 0.012<br>(0.003) |
| HER2+<sup>b</sup> (%) | 10.20 | 10.19 | 6.12 | 4.71 | 0.018<br>(0.006) |
| ER+ or PR+, HER2- | 89.32 | 88.45 | 93.00 | 94.82 | 0.014<br>(0.008) |
| TNBC<sup>c</sup> (%) | 1.20 | 0.82 | 0.40 | 0.40 | 0.373<br>(0.897) |
| Node positive (%) | 49.47 | 46.32 | 47.90 | 40.73 | 0.039<br>(0.066) |
| Stage > 1 (%) | 73.48 | 65.86 | 67.66 | 57.85 | <0.001<br>(0.001) |
| Tumor size > 20mm (%) | 67.80 | 56.13 | 59.86 | 46.76 | <0.001<br>(<0.001) |
| Proliferation score<sup>d</sup> (mean) | 8.87 | 8.80 | 8.55 | 8.50 | <0.001<br>(<0.001) |
| Recurrence score<sup>d</sup> (mean) | 60.17 | 46.99 | 37.63 | 30.83 | <0.001<br>(<0.001) |
| MammaPrint® High-risk (%) | 27.80 | 13.61 | 4.75 | 2.03 | <0.001<br>(<0.001) |
| Oncotype DX® (mean) | 36.44 | 36.02 | 34.06 | 27.67 | <0.001<br>(<0.001) |
| Oncotype DX® High-risk (%) | 56.95 | 54.42 | 54.58 | 35.93 | <0.001<br>(<0.001) |
| Somatic mutations (%) |  |  |  |  |  |
| TP53 | 15.93 | 15.31 | 8.47 | 5.76 | <0.001<br>(<0.001) |
| PIK3CA | 36.95 | 45.58 | 52.54 | 64.75 | <0.001<br>(<0.001) |
| CBFB | 3.39 | 6.80 | 6.10 | 9.49 | 0.004<br>(0.006) |

<sup>a</sup> ER positive by immunohistochemistry; <sup>b</sup> HER2 positive by IHC or FISH; <sup>c</sup> Triple-negative by IHC; <sup>d</sup> Proliferation and Recurrence score by PAM50 genes

### Survival analysis in the combined cohort for pLumA

**Figure 2** shows Kaplan-Meier plots of overall survival for Luminal A cases in the combined cohort stratified into quartiles by Luminal A proportions. There was a statistically significant difference (*P* < 2 × 10^−6^) between Q1 and Q4 cases identified by pLumA. Median overall survival times for Q4 versus Q1 were 232 and 139 months, respectively, and Q1 cases had an estimated 10-year survival probability of only 0.58 (95% CI: 0.51-0.65), compared to 0.76 (95% CI: 0.69-0.82) for Q4. **Table 2** shows the hazard ratios for overall mortality in the combined cohort stratified into quartiles by Luminal A purity. In unadjusted models, the mortality risk for Q1 cases was more than double the risk for Q4 cases. In models adjusted for age, tumor stage and size, hazard ratios were statistically significant but generally lower, as expected due to adjustment for some but not all mediating risk factors.

**Table 2.** Hazard ratios for overall survival from Cox proportional hazards modeling, according to quartiles for Luminal A purity; Luminal A cases in the combined TCGA-METABRIC cohort.

| pLumA | Q1 (admixed) | Q2 | Q3 | Q4 (purest) |  |
| --- | --- | --- | --- | --- | --- |
|  | HR<br>(95% CI) | HR<br>(95% CI) | HR<br>(95% CI) | HR<br>(95% CI) | *P* trend |
| Unadjusted | 2.08<br>(1.58-2.73) | 1.61<br>(1.21-2.14) | 1.36<br>(1.01-1.83) | 1.00<br>- | 5.7x10<sup>-8</sup> |
| Adjusted<sup>a</sup> | 1.70<br>(1.24-2.31) | 1.32<br>(0.95-1.82) | 1.13<br>(0.81-1.57) | 1.00<br>- | 3.1x10<sup>-4</sup> |

<sup>a</sup> Adjusted for age, tumor size and stage

**Figure 2.**
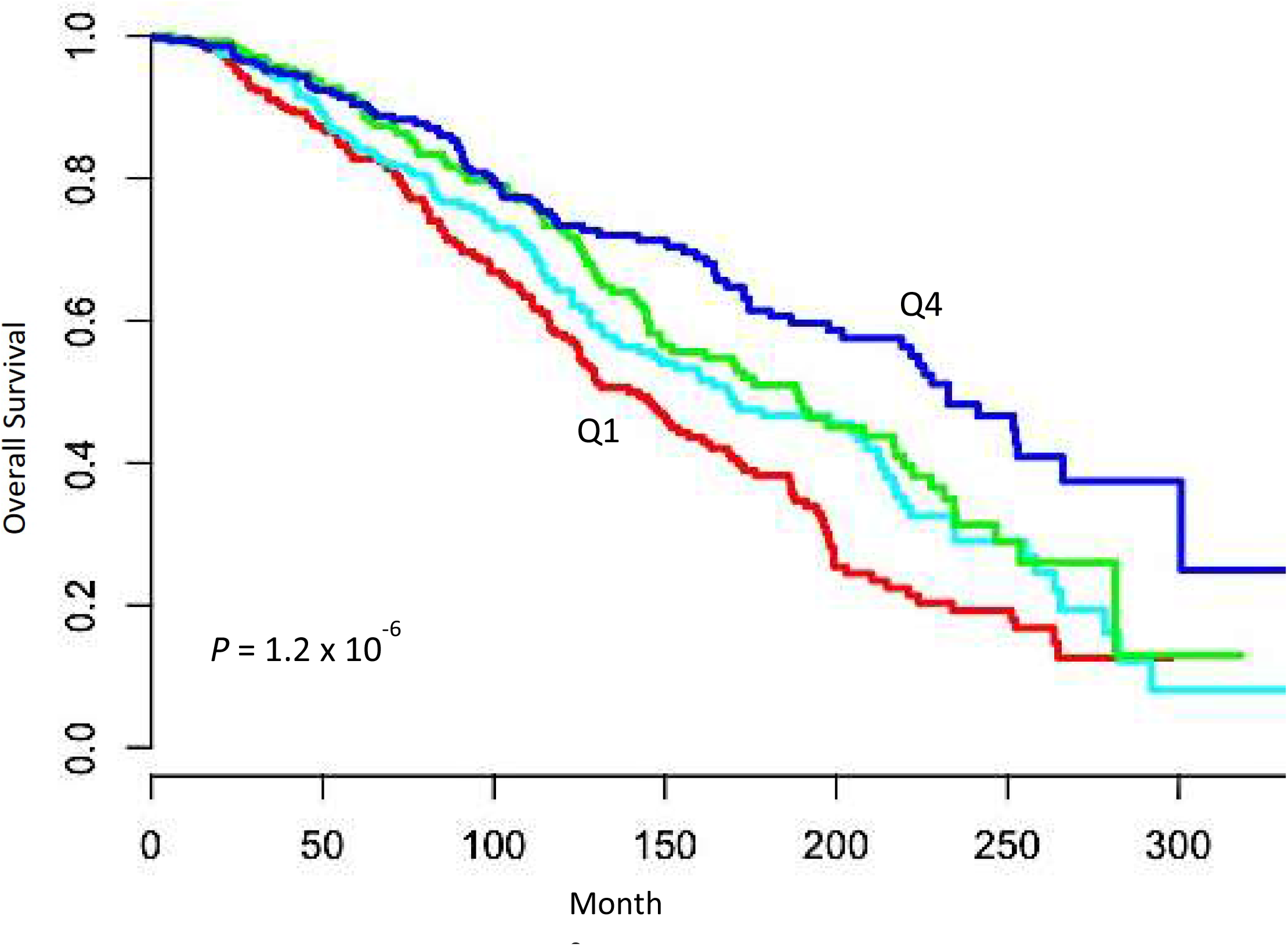
Overall survival of Luminal A breast cancer cases in combined TCGA and METABRIC cohorts, stratified by quartile of transcriptome-based purity measured as pLumA. Q1 = red, Q2 = cyan, Q3 = green, Q4 = blue.

Shannon entropy and pLumA were essentially equivalent as measures of subtype purity, based on associations with clinical/molecular characteristics and survival (**Table S2, Figure S1**). However, as an overall metric of similarity in ssNMF proportions across subtypes, entropy cannot be used to discern which specific subtypes are indicated in the admixture. DRC had weaker associations with tumor characteristics and survival (**Table S3, Figure S1**).

### Comparison of pure Luminal A versus cases with a specific alternate subtype

**Table 3** presents the differences in clinical and molecular characteristics between pure LumA cases and those with a specific alternate subtype. Compared to pure LumA cases, those with predominant LumB admixture were on average 2.6 years older and less likely to be PR positive, with no significant difference in ER or HER2 status. Although these admixed cases showed only small, non-significant increases in node positivity and stage, they were significantly more likely to have tumor size greater than 20mm, and had higher scores for proliferation, recurrence, MammaPrint and Oncotype DX gene expression. **Table 3** also shows that LumB admixed cases had a slightly higher prevalence of *TP53* mutation and lower prevalence of mutated *PIK3CA* and *CBFB*.

**Table 3.** Comparison of purest Luminal A breast cancers in the combined cohort (TCGA, METABRIC), to Luminal A cases exclusively in the highest quartile for admixture with an alternate subtype. (*P* value vs. pure LumA in parentheses)

|  | LumA eQ4<br>n = 275 | LumB eQ4<br>n = 182 | HER2 eQ4<br>n = 157 | Basal eQ4<br>n = 219 |
| --- | --- | --- | --- | --- |
| Age (years, mean) | 60.65<br>- | 63.20<br>(0.036) | 63.68<br>(0.012) | 57.94<br>(0.020) |
| ER+<sup>a</sup> (%) | 97.05<br>- | 94.91<br>(0.315) | 92.99<br>(0.052) | 95.43<br>(0.345) |
| PR+ (%) | 84.81<br>- | 76.37<br>(0.027) | 75.00<br>(0.015) | 83.28<br>(0.711) |
| HER2+<sup>b</sup> (%) | 4.36<br>- | 5.49<br>(0.657) | 12.73<br>(0.002) | 10.86<br>(0.005) |
| ER+ or PR+, HER2- | 94.51<br>- | 94.12<br>(0.838) | 86.13<br>(0.003) | 86.62<br>(0.004) |
| TNBC<sup>c</sup> (%) | 0.00<br>- | 0.36<br>(0.399) | 2.92<br>(0.006) | 0.45<br>(0.444) |
| Node positive (%) | 41.25<br>- | 46.24<br>(0.290) | 44.89<br>(0.543) | 51.22<br>(0.029) |
| Stage > 1 (%) | 58.02<br>- | 63.40<br>(0.329) | 70.15<br>(0.017) | 73.93<br>($<0.001$) |
| Tumor size > 20mm (%) | 46.89<br>- | 57.69<br>(0.028) | 68.15<br>($<0.001$) | 63.47<br>($<0.001$) |
| Proliferation score<sup>d</sup> (mean) | 8.52<br>- | 8.91<br>($<0.001$) | 8.79<br>($<0.001$) | 8.36<br>(0.041) |
| Recurrence score<sup>d</sup> (mean) | 30.68<br>- | 59.04<br>($<0.001$) | 59.63<br>($<0.001$) | 52.43<br>($<0.001$) |
| MammaPrint® High-risk (%) | 1.82<br>- | 13.19<br>($<0.001$) | 15.29<br>($<0.001$) | 15.98<br>($<0.001$) |
| Oncotype DX® (mean) | 27.22<br>- | 29.36<br>(0.054) | 36.42<br>($<0.001$) | 43.45<br>($<0.001$) |
| Oncotype DX® High-risk (%) | 35.27<br>- | 35.16<br>(1.000) | 63.06<br>($<0.001$) | 79.45<br>($<0.001$) |
| Somatic mutations (%) |  |  |  |  |
| TP53 | 5.82<br>- | 9.89<br>(0.144) | 21.02<br>($<0.001$) | 7.76<br>(0.469) |
| PIK3CA | 63.27<br>- | 35.16<br>($<0.001$) | 47.13<br>(0.001) | 34.25<br>($<0.001$) |
| CBFB | 8.73<br>- | 3.85<br>(0.056) | 10.83<br>(0.497) | 2.74<br>(0.007) |

<sup>a</sup> ER positive by immunohistochemistry; <sup>b</sup> HER2 positive by IHC or FISH; <sup>c</sup> Triple-negative by IHC; <sup>d</sup> Proliferation and Risk of Recurrence score based on PAM50 genes

Cases with predominant HER2 admixture, compared to pure cases, were on average older by three years, less likely to be ER or PR positive, and nearly three-fold more likely to be HER2 positive (**Table 3**). HER2 admixture, rather than Basal admixture as might be expected, accounted for nearly all triple negative cases in the combined cohort. HER2 admixed cases were associated with higher stage and tumor size, and higher scores for proliferation, recurrence, MammaPrint and Oncotype DX gene expression. Finally, these cases were three-fold more likely than pure ones to have *TP53* mutations while fewer had mutated *PIK3CA*.

In contrast to the other alternate subtypes, cases with predominant Basal admixture were on average 2.7 years younger than pure LumA, were more likely to be HER2 positive, and showed no significant differences in ER or PR status as determined by immunohistochemistry (**Table 3**). Notably, fewer cases with Basal admixture adhered to a surrogate clinical definition of LumA subtype (ER+ or PR+ and HER2−) than pure Luminal A cases. Triple negative cases were too rare to allow comparison. Basal admixed cases were more likely to have positive nodes, higher stage, and larger tumor size. Basal admixed cases had a lower mean proliferation score but higher mean PAM50 recurrence score. Basal admixture was associated with substantially higher Oncotype DX risk scores and higher likelihood of being in the high-risk category by both MammaPrint and Oncotype DX risk stratification. Finally, Basal admixed cases had no significant difference in *TP53* mutations, but substantially lower prevalence of *PIK3CA* and *CBFB* mutations. **Figure 3** shows mean *EGFR* expression, a canonical marker for the basal phenotype, cross-classified by pBasal and pLumA quartiles. *EGFR* expression increased within each pBasal quartile regardless of pLumA level and was highest in the cases that were most admixed, that is those where both pBasal and pLumA were high.

**Figure 3.**
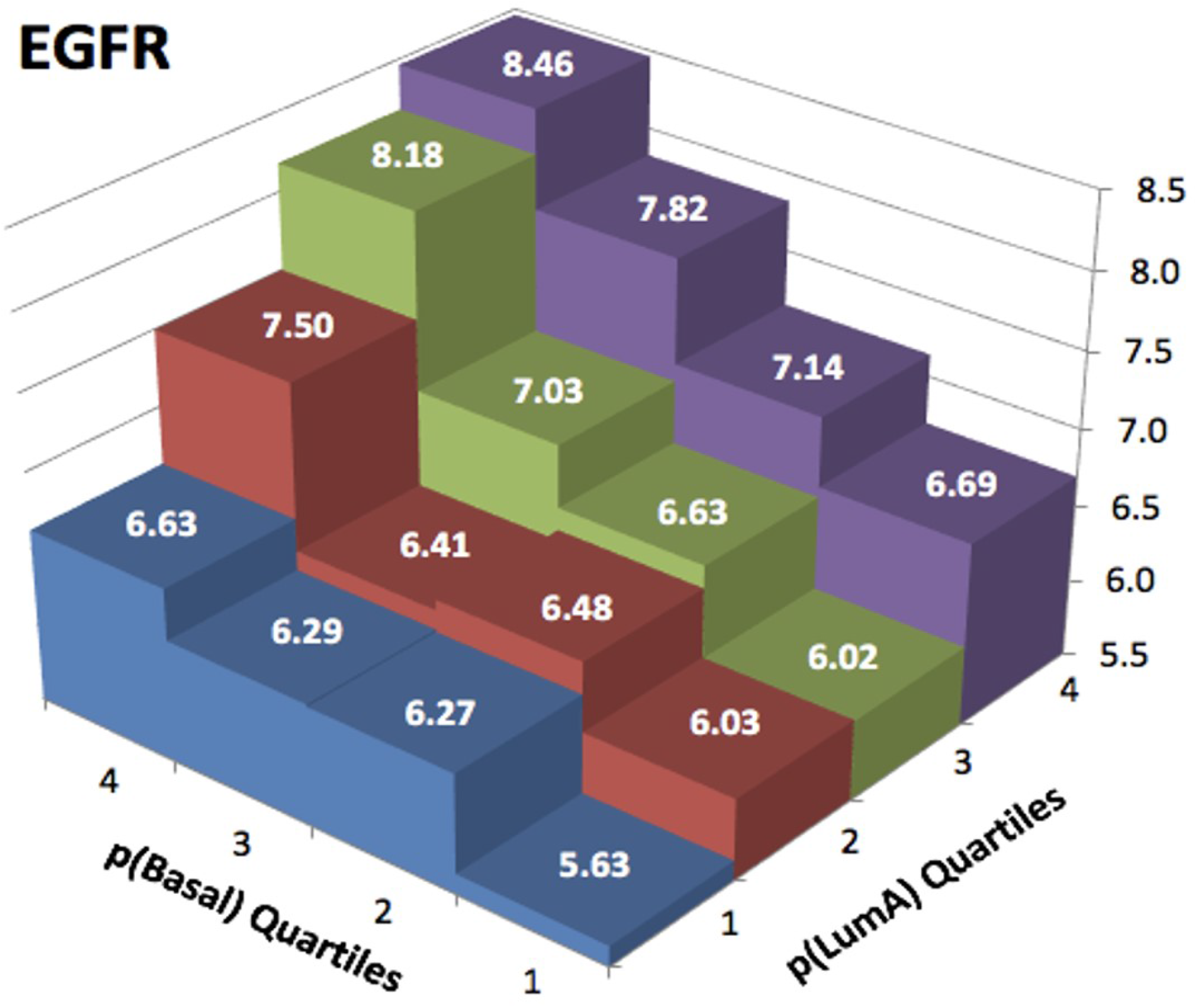
Mean expression of EGFR, a basal-associated gene, increases within each pLumA quartile as pBasal increases.

**Figure 4** shows Kaplan-Meier plots of overall survival for pure LumA, and cases with predominant LumB, HER2, and Basal admixture. There was a significant difference between pure LumA and LumB admixed cases (*P* = 0.030), and a more pronounced difference when the admixture was with HER2 subtype (*P* < 0.001). However, there was essentially no difference in survival between pure LumA versus Basal admixed cases (*P* = 0.515); notably, survival for the latter cases was actually equivalent or slightly better than pure LumA before crossing over to worse after 10 years. The median survival times for pure LumA, LumB-admixed, HER2-admixed, and Basal-admixed cases were 228, 169, 190 and 161 months, respectively. The corresponding 10-year survival probabilities were 0.72 (95% CI: 0.65-0.79), 0.65 (95% CI: 0.56-0.74), 0.66 (95% CI: 0.58-0.82) and 0.64 (95% CI: 0.53-0.74), respectively.

**Figure 4.**
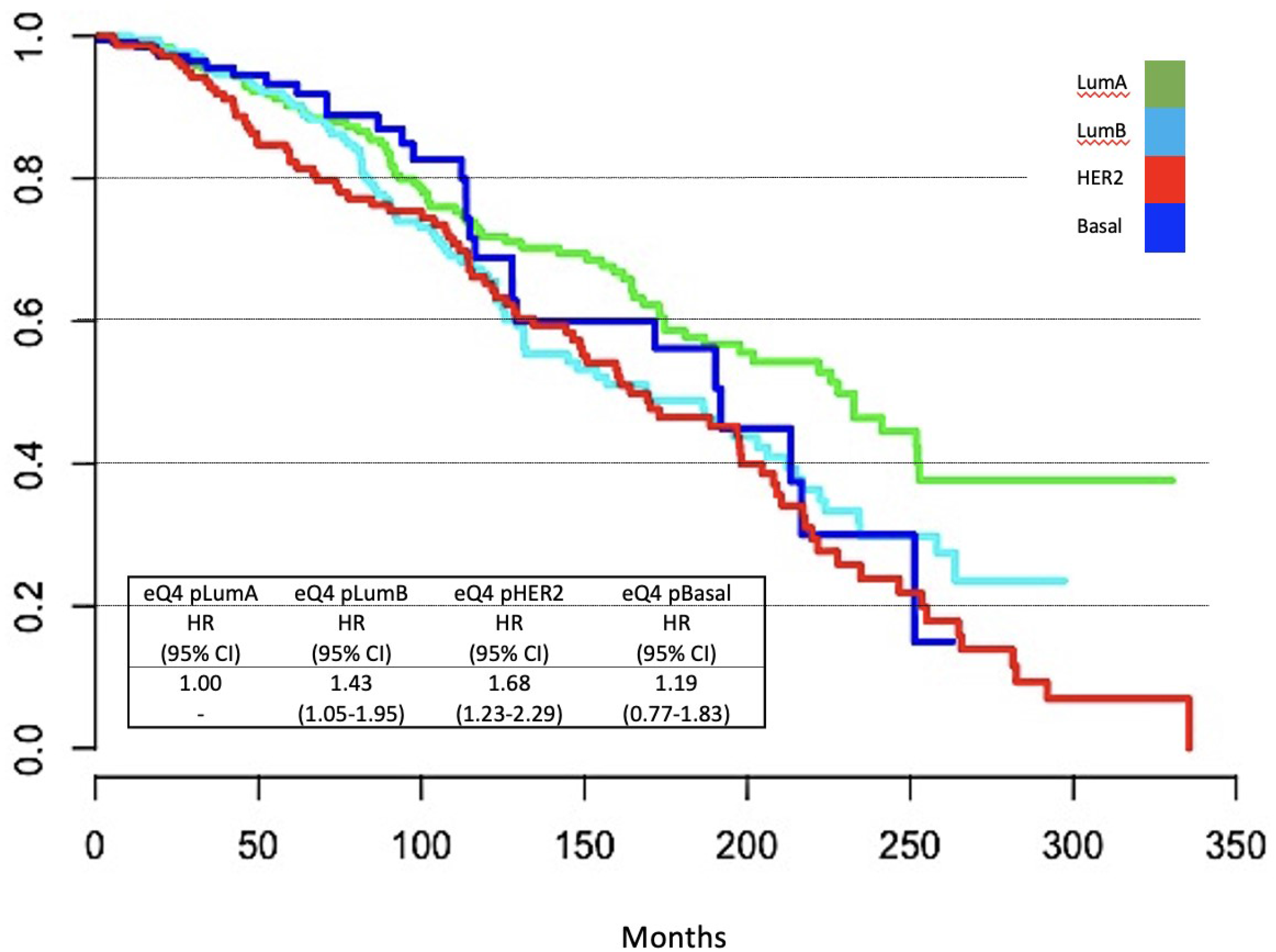
Comparison of overall survival of purest Luminal A breast cancer cases in combined TCGA and METABRIC cohorts, to Luminal A cases in the exclusive highest quartile for admixture with an alternate subtype.

**Table 4** shows the results of Cox-proportional hazards models for overall mortality of pure LumA cases versus those admixed with other subtypes. The hazard ratios for LumB-admixed, HER2-admixed, and Basal-admixed cases relative to pure LumA were 1.43, (*P* = 0.025), 1.68 (*P* = 0.001), and 1.19 (*P* = 0.424) respectively. With adjustment for age, tumor stage and size, the hazard ratios for respective categories decreased to 1.13 (*P* = 0.491), 1.27 (*P* = 0.180) and 0.89 (*P* = 0.639).

**Table 4.**
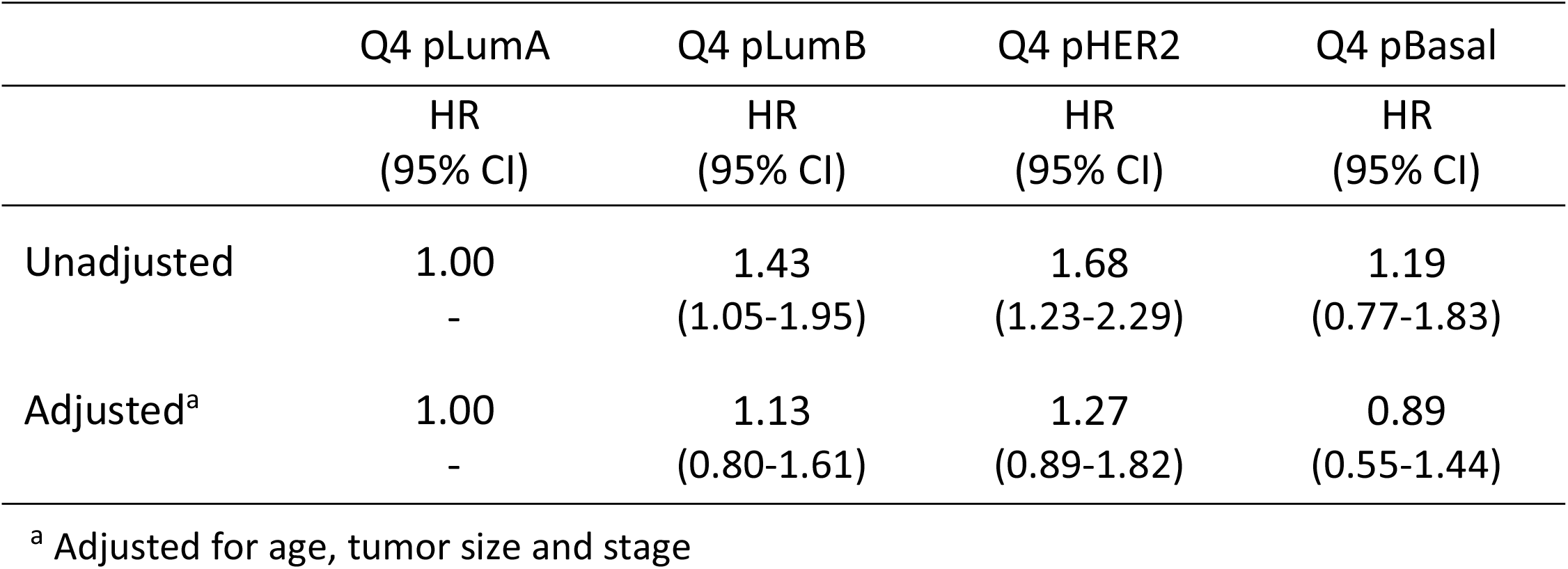
Hazard ratios for overall survival from Cox proportional hazards modeling, comparing Luminal A cases exclusively in the highest quartile for purity (referent) versus Luminal A cases exclusively within the highest quartile for admixture with an alternate subtype; combined TCGA-METABRIC cohort.

We repeated analyses in TCGA and METABRIC separately to check for consistency and evaluate variables unique to each cohort. The associations between pNMF and clinical characteristics were generally similar for the two cohorts, although trends were weaker in TCGA (**Tables S4 and S5**). Data available only in METABRIC showed that cases with low pLumA were substantially more likely to be high-grade and have HER2 copy number gain. In both cohorts, low pLumA was associated with significantly increased risk of mortality compared to more pure cases (**Figure S2**), with hazard ratios of 1.65 (95% CI: 1.28-2.14, P < 0.001) and 1.91 (95% CI: 1.08-3.63, P = 0.002) for METABRIC and TCGA, respectively.

In TCGA we observed no association between pLumA and the number of subclone populations estimated by PyClone, and no significant differences between pure and admixed cases when compared by closest alternate subtype (**Figure S3A and B**). In contrast, we observed predicted relationships between admixture and IntClust grouping in the METABRIC cohort; 97% of the purest LumA cases were classified within IntClust groups 3, 4, 7 and 8, which were previously associated with LumA tumors, whereas the more admixed cases had more diverse distribution (**Figure 5A**), indicating that the transcriptomically admixed tumors had some molecular characteristics linked to alternate subtypes. **Figure 5B** shows the proportions within each Integrative Cluster, of METABRIC Luminal A cases belonging exclusively to the highest quartile for alternate subtypes. Cases with predominant HER2 admixture were enriched in groups 5 and 8, which are typically associated with the HER2 subtype, but no admixed Basal cases were assigned to IntClust 10, which is associated with triple-negative breast cancer.

**Figure 5.**
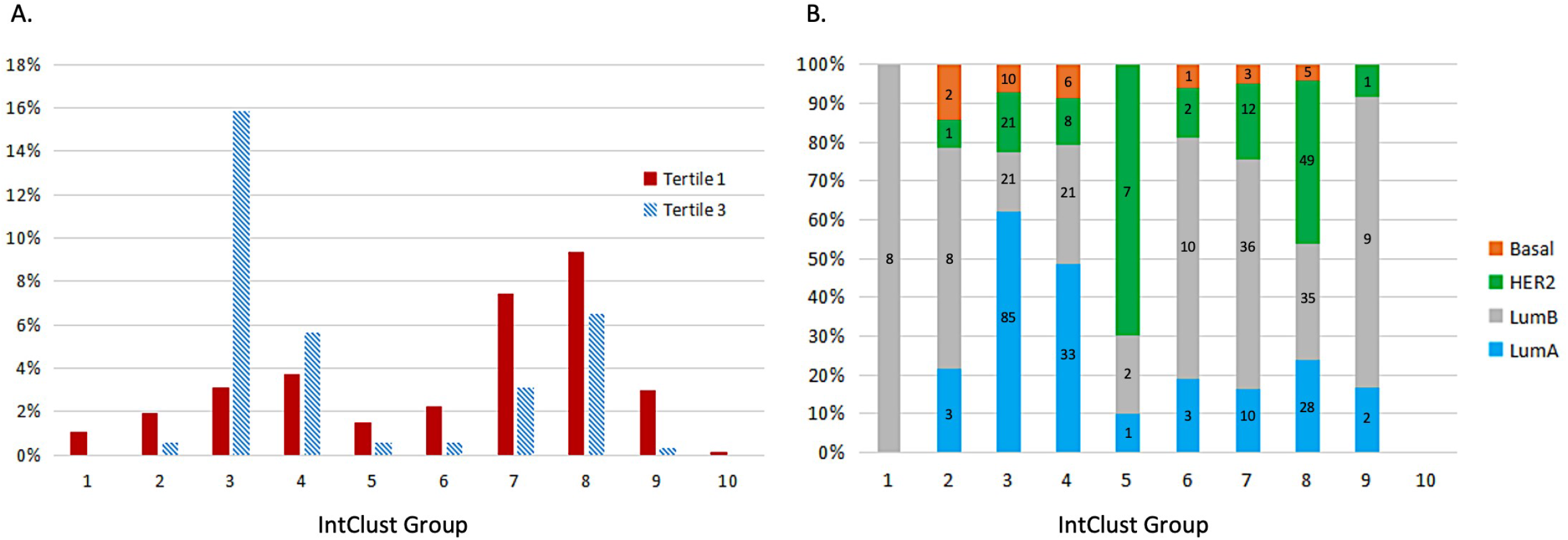
**5A** shows the distribution of METABRIC Luminal A cases across lntClust groups, comparing cases in the highest tertilefor plumA with those in the lowest tertile. **5B** shows the proportions within each Integrative Cluster, of METABRIC Luminal A cases belonging exclusively to the highest (purest) quartile for plumA, plumB, pBasal and pHER2. Counts are shown in each vertical bar.

## DISCUSSION

In this analysis, we demonstrate that semi-supervised non-negative matrix factorization enabled us to measure the degree of adherence of an individual breast cancer case to each of the four major intrinsic subtypes based on its whole transcriptomic profile, thus providing a novel way to evaluate the relationship of subtype purity to tumor characteristics and behavior. We found that cases assigned by PAM50 as Luminal A exhibited a wide range of adherence to LumA purity, and that higher purity was strongly associated with numerous clinical and molecular features linked to better prognosis. Indeed, cases within the highest quartile of adherence to the LumA subtype had less than half the mortality risk of cases within the lowest quartile. We also found that LumA cases whose closest alternative subtype was either LumB or HER2 had tumor features consistent with those subtypes, and survival that was significantly lower than pure LumA cases. We note that survival for pure LumA versus the admixed cases did not diverge until at least three years of follow-up, suggesting that subtype admixture could explain the established observation that while hormone-positive breast cancer patients have better initial survival, a subset is more susceptible to late recurrence [8; 14].

Unexpectedly, LumA cancers with the highest resemblance to the Basal subtype did not consistently display features typically associated with Basal-like (or triple negative) breast cancers. These patients, as predicted, were younger than those with pure LumA and were more likely to have larger tumors and nodal metastasis. However, cases with high Basal admixture had the same likelihood of ER and PR positivity, suggesting that coexisting basal components have a luminal phenotype. Additionally, basal-admixed cases had lower proliferation compared to pure LumA, and overall survival was statistically indistinguishable. Paradoxically, the highest expression of EGFR, a marker classically associated with basal phenotype and poor prognosis, was observed in cases with the highest level of both Basal and Luminal A transcriptomic profile. These data suggest that cases that are predominantly Luminal A but have Basal-like admixture are quite distinct from typical Luminal A or Basal cases, based on both molecular and clinical characteristics. Furthermore, the discordance between various gene-expression based scores for risk of recurrence and patient survival, which was observed only for Basal-admixed cases, could indicate that the risk estimation is less accurate in this subgroup. Due to such counterintuitive findings, we speculated that these cases might represent admixture with the Luminal Androgen Receptor Subtype (LARS) of triple-negative breast cancer, which displays significant ER and PR expression, but we found no associations between LARS signature gene panels and degree of basal admixture, and only rare occurrence of triple negative status [15; 16].

These results support the conclusion that bulk sampling of tumors for genomic analysis can provide an opportunity to expose intratumoral heterogeneity, such as intrinsic subtype admixture.[17] While our approach expands the notion of how substantial genomic diversity within LumA cases actually is, the scale at which this diversity manifests itself is not immediately clear. We can envision three possibilities: first, that all cells within a tumor express the same admixed profile; second, that subtype adherence varies from cell to neighboring cell; or third, that multiclonality leads to larger clusters of cells expressing divergent profiles. The robustness of the PAM50 classifier for predicting clinical outcomes implies that each subtype represents a favorable genomic profile or pathway for subclonal expansion, thus favoring the third hypothesis. The true nature of this subtype admixture could be elucidated by emerging but relatively costly methods such as single-cell RNAseq or high-dimensional spatial profiling [18].

Intratumor heterogeneity involving breast cancer subtypes has been indirectly implicated to explain differences in outcomes when PAM50 and IHC subtype classifications are discordant in a primary tumor [19], when subtypes are discordant between a synchronous primary tumor and metastases [20], or when comparing treatment response for HER2 cases with or without ER positivity [21]. Moreover, the ASCO/CAP criteria for subtype classification only require ER expression greater than 1% of cells, or HER2 overexpression in greater than 10%; and for in situ hybridization-based assays, a count in 20 cells is sufficient with no percentage threshold having been established [22; 23]. Compared to tumors with higher levels of ER expression, tumors with low levels of ER-positivity (1-10%) are more likely to be classified as basal-like and less likely to be responsive to endocrine therapy [24]. Similarly, HER2-positive cases with a smaller proportion of amplified cells are less responsive to HER2-targeted therapy [21]. Examples of more direct characterization of intratumor heterogeneity include the discovery of lumino-basal cells in ER-positive tumors, [25], discordant multiregional DNA sequencing, [26] discordant IHC staining across tissue microarray cores or whole slides, [27] single cell HER2 FISH analysis, [28] and early results from high-dimensional spatial profiling [18; 29].

We previously reported that a simpler subtype admixture metric, based only on PAM50 genes, was also associated with divergent tumor characteristics and behavior among LumA cases [1]. Camp and coworkers used principal components analysis to derive quantitative metrics based on PAM50 gene expression and found that it could also uncover subgroups with survival and treatment response that was independent of assigned subtype [30]. However, the current method used the entire transcriptome to compute a continuous rather than categorical metric that demonstrates stronger associations and provides construct validity by showing that admixed cases have features resembling their closest alternate subtype. Other strengths of this study include the large number of genes with expression levels available for analysis, the large size of the combined cohorts, and the similarity of the results between the two cohorts. However, there is a need for further validation of our findings in additional independent populations.

In summary, we have developed a metric based on whole transcriptome data that can stratify LumA cancers based on subtype purity and thus provide information that is potentially predictive with respect to prognosis and treatment response. Extensions of this work could include examination of the metagenes resulting from NMF to discover pathways that are up- or down-regulated by subtype admixture, and identification of smaller gene sets for enhanced clinical prediction modeling. In addition, our method can be used to test the association of admixture with treatment response, for any assigned subtype.

## Data Availability

Data used is publicly available from the original sources.

https://portal.gdc.cancer.gov/

https://www.mercuriolab.umassmed.edu/metabric

## Acknowledgments

The authors thank Yash Dharmamer and Dan Zhao for their help with this work. They also express their gratitude to the research teams who developed the TCGA and METABRIC resources and made them available to the scientific community, and to the patients whose data and biospecimens were donated for these cohorts.

---

**Table S1.** Key features of the TCGA and METABRIC datasets

| **Table S1.** Key features of the TCGA and METABRIC datasets |  |  |  |
| --- | --- | --- | --- |
| *Feature* | *TCGA-BRCA* | *METABRIC* | *Notes* |
| All genes | 20,532 | 17,814 | 15,747 genes overlap |
| PAM50 genes | 50 | 47 | 47 genes overlap |
| Genomic technology | Illumina HiSeq (mRNA) | Illumina HT-12v3 (mRNA); Affymetrix SNP 6.0 array (CNA) |  |
| No. of cases | 1,081 | 1,980 | Luminal A: 505 (TCGA), 674 (METABRIC) |
| Normalization done at source | Median centering | Median centering and log transform |  |
| Normalization done by authors | Log transform | Linear transform to match mean and variance to TCGA cohort for each gene |  |

**Table S2.** Characteristics of Luminal A breast cancers in the combined cohort (TCGA, METABRIC), stratified by quartile of negative Shannon’s Entropy subtype purity based on transcriptome

|  | **Q1**<br>n = 295 | **Q2**<br>n = 294 | **Q3**<br>n = 295 | **Q4**<br>n = 295 | **P, Q1 vs Q4**<br>**(p trend)** |
| --- | --- | --- | --- | --- | --- |
| Age (mean) | 63.06 | 62.04 | 60.35 | 60.03 | 0.006<br>(0.002) |
| ER+<sup>a</sup> (%) | 97.23 | 97.93 | 98.25 | 98.93 | 0.222<br>(0.186) |
| PR+ (%) | 78.32 | 80.76 | 85.02 | 85.22 | 0.043<br>(0.013) |
| HER2+<sup>b</sup> (%) | 10.82 | 8.46 | 7.63 | 4.31 | 0.005<br>(0.007) |
| ER+ or PR+, HER2- | 88.62 | 90.35 | 91.42 | 95.22 | 0.004<br>(0.008) |
| TNBC<sup>c</sup> (%) | 1.20 | 0.86 | 0.40 | 0.40 | 0.373<br>(0.902) |
| Node positive (%) | 49.64 | 44.92 | 45.07 | 44.96 | 0.322<br>(0.298) |
| Stage > 1 (%) | 72.23 | 66.54 | 64.56 | 62.03 | 0.006<br>(0.014) |
| Tumor size > 20mm (%) | 63.73 | 58.64 | 57.97 | 50.17 | 0.001<br>(0.001) |
| Proliferation score<sup>d</sup> (mean) | 8.94 | 8.78 | 8.58 | 8.42 | <0.001<br>(<0.001) |
| Recurrence score<sup>d</sup> (mean) | 58.97 | 48.31 | 38.97 | 29.38 | <0.001<br>(<0.001) |
| Mammaprint® High-risk (%) | 24.40 | 14.97 | 7.80 | 1.02 | <0.001<br>(<0.001) |
| Oncotype DX® (mean) | 35.89 | 34.78 | 35.47 | 28.05 | <0.001<br>(<0.001) |
| Oncotype DX® High-risk (%) | 54.92 | 50.00 | 60.00 | 36.95 | <0.001<br>(<0.001) |
| Somatic mutations (%) |  |  |  |  |  |
| TP53 | 17.29 | 15.93 | 7.14 | 5.08 | <0.001<br>(<0.001) |
| PIK3CA | 44.41 | 42.32 | 50.34 | 62.71 | <0.001<br>(<0.001) |
| CBFB | 3.37 | 9.15 | 3.40 | 9.49 | 0.004<br>(0.074) |

<sup>a</sup> ER positive by immunohistochemistry; <sup>b</sup> HER2 positive by IHC or FISH; <sup>c</sup> Triple-negative by IHC; <sup>d</sup> Proliferation and Recurrence score by PAM50 genes

**Table S3.** Characteristics of Luminal A breast cancers in the combined cohort (TCGA, METABRIC), stratified by quartile of -DRC subtype purity based on transcriptome

|  | **Q1**<br>n = 295 | **Q2**<br>n = 294 | **Q3**<br>n = 295 | **Q4**<br>n = 295 | **P, Q1 vs Q4**<br>**(p trend)** |
| --- | --- | --- | --- | --- | --- |
| Age (mean) | 62.32 | 61.98 | 60.78 | 60.40 | 0.072<br>(0.039) |
| ER+<sup>a</sup> (%) | 97.60 | 98.23 | 98.60 | 97.92 | 1.000<br>(1.000) |
| PR+ (%) | 71.13 | 83.80 | 86.90 | 86.59 | <0.001<br>(<0.001) |
| HER2+<sup>b</sup> (%) | 10.08 | 8.67 | 5.81 | 6.75 | 0.183<br>(0.190) |
| ER+ or PR+, HER2- | 89.86 | 88.89 | 94.05 | 92.80 | 0.241<br>(0.079) |
| TNBC<sup>c</sup> (%) | 1.45 | 0.85 | 0.00 | 0.40 | 0.373<br>(0.084) |
| Node positive (%) | 47.90 | 44.57 | 45.75 | 46.26 | 0.741<br>(0.769) |
| Stage > 1 (%) | 67.03 | 66.67 | 62.11 | 69.06 | 0.659<br>(0.900) |
| Tumor size > 20mm (%) | 62.03 | 60.27 | 53.22 | 55.10 | 0.112<br>(0.032) |
| Proliferation score<sup>d</sup> (mean) | 9.06 | 8.84 | 8.64 | 8.18 | <0.001<br>(<0.001) |
| Recurrence score<sup>d</sup> (mean) | 69.17 | 51.09 | 35.86 | 19.45 | <0.001<br>(<0.001) |
| Mammaprint® High-risk (%) | 26.45 | 12.93 | 6.78 | 2.03 | <0.001<br>(<0.001) |
| Oncotype DX® (mean) | 33.85 | 37.90 | 32.38 | 30.08 | <0.001<br>(<0.001) |
| Oncotype DX® High-risk (%) | 51.86 | 57.14 | 47.12 | 45.76 | <0.001<br>(<0.001) |
| Somatic mutations (%) |  |  |  |  |  |
| TP53 | 17.29 | 12.59 | 10.85 | 4.75 | <0.001<br>(<0.001) |
| PIK3CA | 46.78 | 44.56 | 50.85 | 57.63 | 0.011<br>(0.003) |
| CBFB | 6.10 | 4.75 | 8.16 | 6.78 | 0.867<br>(0.402) |

<sup>a</sup> ER positive by immunohistochemistry; <sup>b</sup> HER2 positive by IHC or FISH; <sup>c</sup> Triple-negative by IHC; <sup>d</sup> Proliferation and Recurrence score by PAM50 genes

**Table S4.**
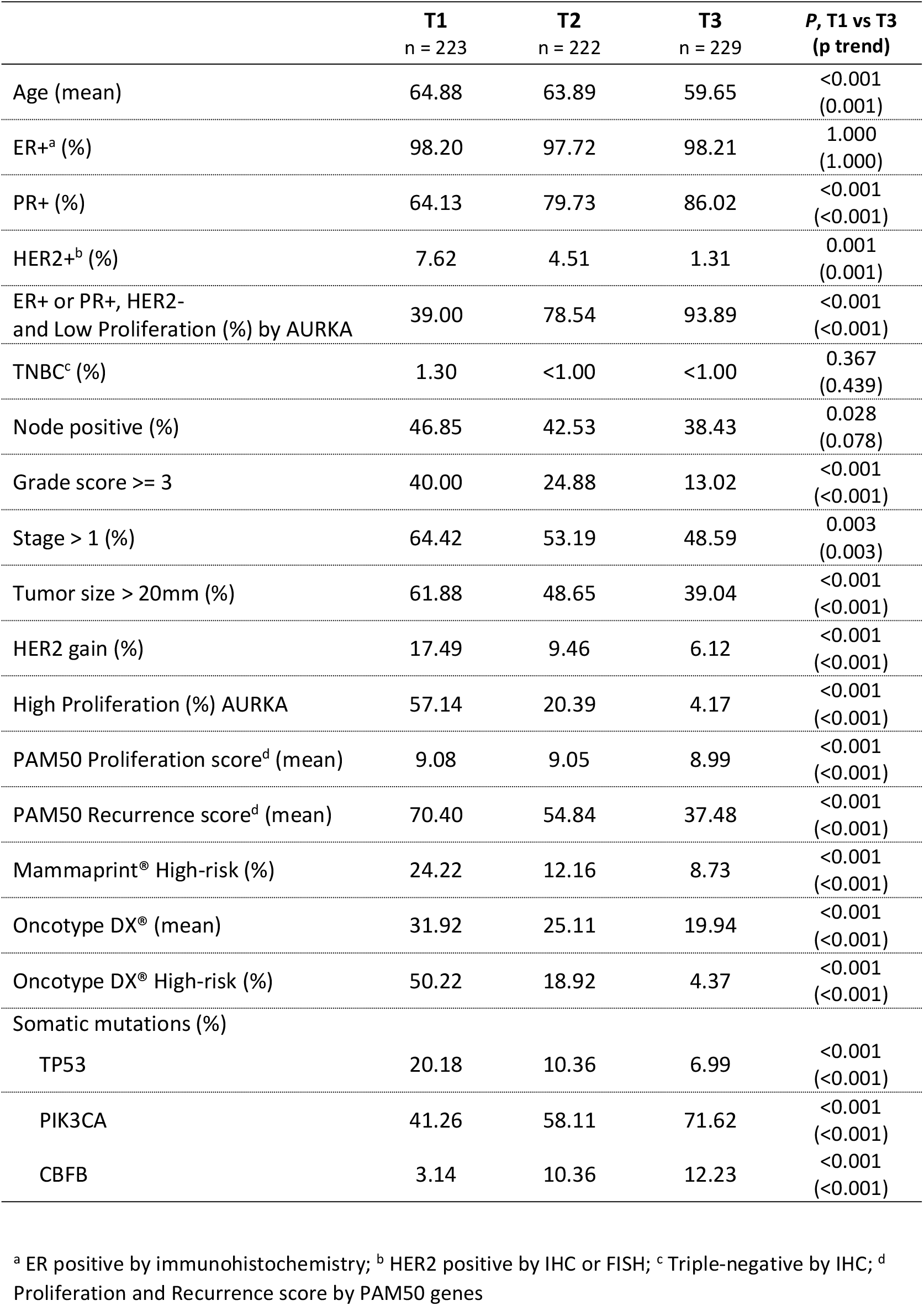
Characteristics of Luminal A breast cancers in the METABRIC cohort, stratified by tertile of pLumA subtype purity based on transcriptome

|  | **T1**<br>n = 223 | **T2**<br>n = 222 | **T3**<br>n = 229 | **P, T1 vs T3**<br>**(p trend)** |
| --- | --- | --- | --- | --- |
| Age (mean) | 64.88 | 63.89 | 59.65 | <0.001<br>(0.001) |
| ER+<sup>a</sup> (%) | 98.20 | 97.72 | 98.21 | 1.000<br>(1.000) |
| PR+ (%) | 64.13 | 79.73 | 86.02 | <0.001<br>(<0.001) |
| HER2+<sup>b</sup> (%) | 7.62 | 4.51 | 1.31 | 0.001<br>(0.001) |
| ER+ or PR+, HER2-<br>and Low Proliferation (%) by AURKA | 39.00 | 78.54 | 93.89 | <0.001<br>(<0.001) |
| TNBC<sup>c</sup> (%) | 1.30 | <1.00 | <1.00 | 0.367<br>(0.439) |
| Node positive (%) | 46.85 | 42.53 | 38.43 | 0.028<br>(0.078) |
| Grade score >= 3 | 40.00 | 24.88 | 13.02 | <0.001<br>(<0.001) |
| Stage > 1 (%) | 64.42 | 53.19 | 48.59 | 0.003<br>(0.003) |
| Tumor size > 20mm (%) | 61.88 | 48.65 | 39.04 | <0.001<br>(<0.001) |
| HER2 gain (%) | 17.49 | 9.46 | 6.12 | <0.001<br>(<0.001) |
| High Proliferation (%) AURKA | 57.14 | 20.39 | 4.17 | <0.001<br>(<0.001) |
| PAM50 Proliferation score<sup>d</sup> (mean) | 9.08 | 9.05 | 8.99 | <0.001<br>(<0.001) |
| PAM50 Recurrence score<sup>d</sup> (mean) | 70.40 | 54.84 | 37.48 | <0.001<br>(<0.001) |
| Mammaprint® High-risk (%) | 24.22 | 12.16 | 8.73 | <0.001<br>(<0.001) |
| Oncotype DX® (mean) | 31.92 | 25.11 | 19.94 | <0.001<br>(<0.001) |
| Oncotype DX® High-risk (%) | 50.22 | 18.92 | 4.37 | <0.001<br>(<0.001) |
| Somatic mutations (%) |  |  |  |  |
| TP53 | 20.18 | 10.36 | 6.99 | <0.001<br>(<0.001) |
| PIK3CA | 41.26 | 58.11 | 71.62 | <0.001<br>(<0.001) |
| CBFB | 3.14 | 10.36 | 12.23 | <0.001<br>(<0.001) |

<sup>a</sup> ER positive by immunohistochemistry; <sup>b</sup> HER2 positive by IHC or FISH; <sup>c</sup> Triple-negative by IHC; <sup>d</sup> Proliferation and Recurrence score by PAM50 genes

**Table S5.** Characteristics of Luminal A breast cancers in the TCGA cohort, stratified by tertile of pLumA subtype purity based on transcriptome

|  | **T1**<br>n = 167 | **T2**<br>n = 166 | **T3**<br>n = 172 | **P, T1 vs T3**<br>**(p trend)** |
| --- | --- | --- | --- | --- |
| Age (mean) | 58.43 | 59.87 | 60.19 | 0.064<br>(0.218) |
| ER+<sup>a</sup> (%) | 97.58 | 98.14 | 98.73 | 1.000<br>(0.699) |
| PR+ (%) | 87.66 | 90.24 | 92.63 | 0.479<br>(0.330) |
| HER2+<sup>b</sup> (%) | 23.63 | 17.43 | 11.12 | 0.014<br>(0.034) |
| ER+ or PR+, HER2- | 76.36 | 80.73 | 88.70 | 0.021<br>(0.041) |
| TNBC<sup>c</sup> (%) | 1.00 | 1.00 | 0.00 | 1.000<br>(1.000) |
| Node positive (%) | 54.49 | 49.65 | 51.28 | 0.265<br>(0.670) |
| Stage > 1 (%) | 79.63 | 79.87 | 75.89 | 0.431<br>(0.315) |
| Tumor size > 20mm (%) | 72.09 | 67.47 | 64.67 | 0.131<br>(0.109) |
| PAM50 Proliferation score<sup>d</sup> (mean) | 8.45 | 8.18 | 7.95 | <0.001<br>(<0.001) |
| PAM50 Recurrence score<sup>d</sup> (mean) | 35.09 | 29.27 | 26.56 | <0.001<br>(<0.001) |
| Mammaprint® High-risk (%) | 19.16 | 10.24 | 5.81 | <0.001<br>(<0.001) |
| Oncotype DX® (mean) | 49.95 | 43.64 | 38.99 | <0.001<br>(<0.001) |
| Oncotype DX® High-risk (%) | 89.22 | 88.55 | 78.49 | <0.001<br>(<0.001) |
| Mutational load (median) | 26.50 | 23.00 | 22.00 | 0.449<br>(0.432) |
| MATH score (mean) | 0.363 | 0.382 | 0.381 | 0.232<br>(0.323) |
| Somatic mutations (%) |  |  |  |  |
| TP53 | 16.17 | 11.45 | 2.33 | <0.001<br>(<0.001) |
| PIK3CA | 31.74 | 41.57 | 47.09 | <0.001<br>(<0.001) |
| CBFB | 2.99 | 3.01 | 4.65 | 0.574<br>(0.640) |

<sup>a</sup> ER positive by immunohistochemistry; <sup>b</sup> HER2 positive by IHC or FISH; <sup>c</sup> Triple-negative by IHC; <sup>d</sup> Proliferation and Recurrence score by PAM50 genes

**Figure S1.**
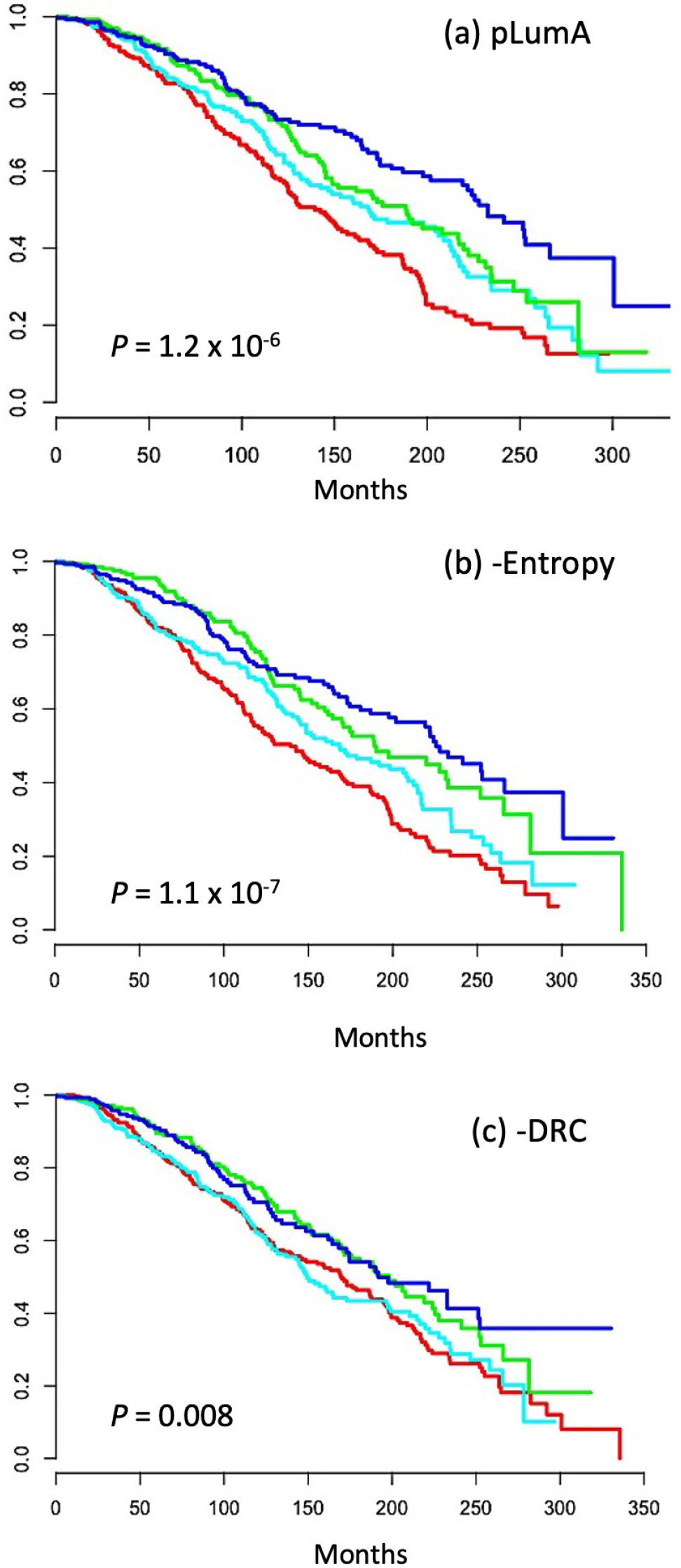
Overall survival of Luminal A breast cancer cases in combined TCGA and METABRIC cohorts, stratified by quartile of transcriptome-based purity measured as (a) pLumA, (b) Shannon’s entropy and, (c) distance ratio criteria (DRC). Q1 = red, Q2 = cyan, Q3 = green, Q4 = blue.

**Figure S2.**
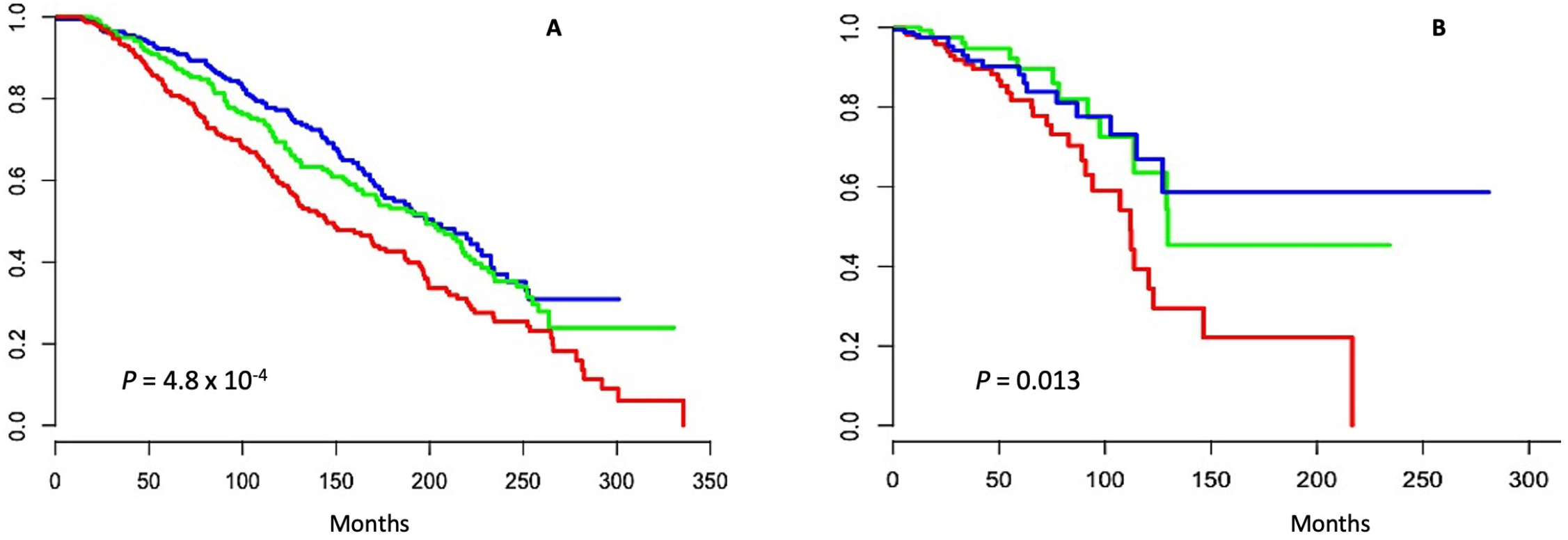
Overall survival of Luminal A breast cancer cases in (a) METABRIC cohort and (b) in TCGA cohort, stratified by tertile of respective transcriptome-based purity measured as pLumA, T1 = red, T2 = green, T3= blue.

**Figure S3.**
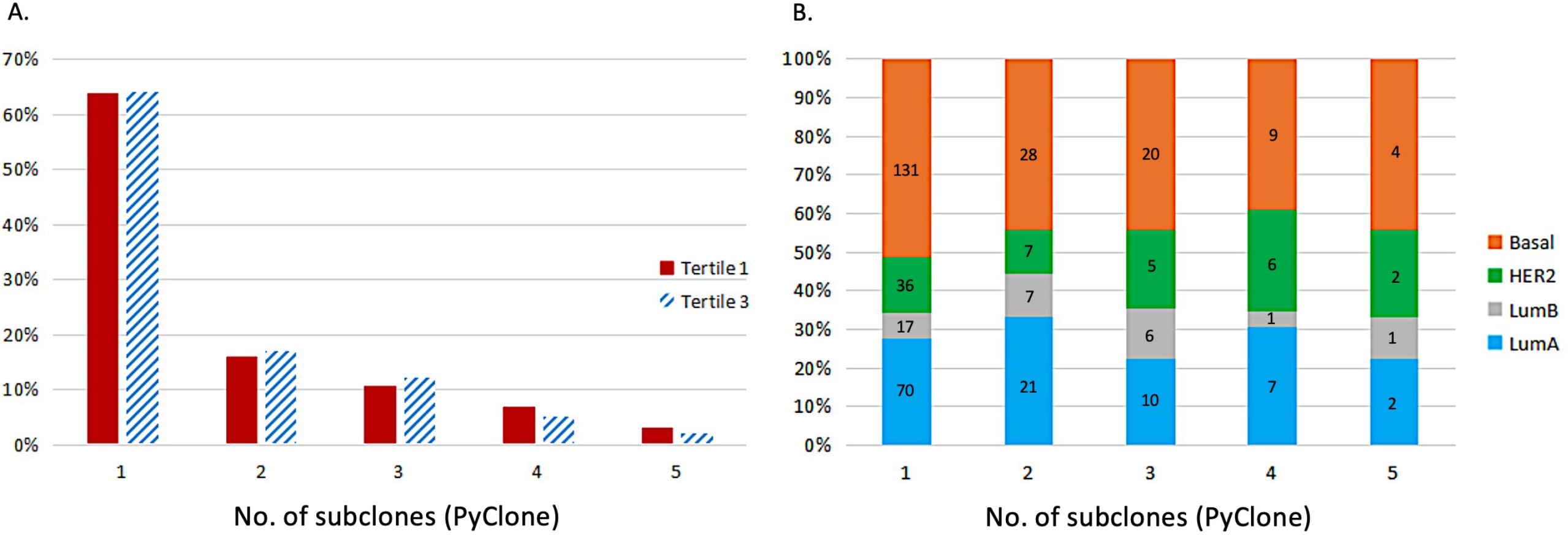
**A**: Number of subclones estimated by PyClone, comparing the highest (purest) tertile for pLumA to the lowest (most admixed) tertile, among Luminal A cases in TCGA. Tl (admixed), T3 (pure). **B**: Distribution of cases within each subclone number based on pure LumA and exclusive alternate subtypes. Counts are shown in each vertical bar. Subtype admixture was not associated with subclone number in either analysis.

